# Inflammation and autoreactivity define a discrete subset of patients with post-acute sequelae of COVID-19, or long-COVID

**DOI:** 10.1101/2021.09.21.21263845

**Authors:** Matthew C. Woodruff, Kevin S. Bonham, Fabliha A. Anam, Tiffany A. Walker, Yusho Ishii, Candice Y. Kaminski, Martin C. Ruunstrom, Alexander D. Truong, Adviteeya N. Dixit, Jenny E. Han, Richard P. Ramonell, Natalie S. Haddad, Mark E. Rudolph, Scott A. Jenks, Arezou Khosroshahi, F. Eun-Hyung Lee, Ignacio Sanz

## Abstract

While significant attention has been paid to the immunologic determinants of disease states associated with COVID-19^1,2^, their contributions to post-acute sequelae of COVID-19 (PASC) remain less clear^3-5^. Due to the wide array of PASC presentations^6^, it is critical to understand if specific features of the disease are associated with discrete immune processes, and whether those processes may be therapeutically targeted. To this end, we performed wide immunologic and serological characterization of patients in the early recovery phase of COVID-19 across a breadth of symptomatic presentations. Using high-parameter proteomics screening and applied machine learning (ML), we identify clear signatures of immunologic activity between PASC patients and uncomplicated recovery, dominated by inflammatory cytokine signaling, neutrophil activity, and markers of cell death. Consistent with disease complexity, heterogeneity in plasma profiling reveals distinct PASC subsets with striking divergence in these ongoing inflammatory processes, here termed plasma quiescent (plaq) and inflammatory (infl) PASC. In addition to elevated inflammatory blood proteomics, inflPASC patients display positive clinical tests of acute inflammation including C-reactive protein and fibrinogen, increased B cell activity with extrafollicular involvement coupled with elevated targeting of viral nucleocapsid protein and clinical autoreactivity. Further, the unique plasma signatures of PASC patients allowed for the creation of refined models with high sensitivity and specificity for the positive identification of inflPASC with a streamlined assessment of 12 blood markers. Additionally, refined ML modeling highlights the unexpected significance of several markers of potential diagnostic or therapeutic use for PASC in general, including the peptide hormone, epiregulin. In all, this work identifies clear biological signatures of PASC with potential diagnostic and therapeutic potential and establishes clear disease subtypes that are both easily identifiable and highly relevant to ongoing efforts in both therapeutic targeting and epidemiological investigation of this highly complex disease.

## Introduction

The COVID-19 pandemic resulting from the emergence of the novel beta-coronavirus SARS-CoV-2 has been characterized by significant heterogeneity in disease manifestations^7^, clinical outcomes^8^, and recovery^9^. A particularly important aspect of those investigations has become an increased focus on patients that, despite resolution of many of the symptoms associated with acute viral infection, experience ongoing complications^10^. These Post-Acute Sequelae of COVID-19 (PASC) range both in manifestation and severity from anosmia to fatigue to joint pain persisting months or even years following the acute phase of disease^11^. Although a continuum of disease has been clearly documented from the acute phase in patient cohorts^12^, the US Center for Disease Control and the World Health Organization recognize PASC diagnosis at 4 and 12 weeks after COVID-19 onset, respectively, to allow for acute phase response recovery^13,14^. While significant effort has generated an expansive collection of immunologic associations across a spectrum of COVID-19 disease courses, their differential resolution and potential contribution in PASC remains less clear^1^.

Reliable immunotypes of severe/critical versus mild/moderate COVID-19 dependent on, or contributing to, a high-inflammation environment have been identified in acute disease^15^. In particular, integration of systems approaches to immune assessment have identified prominent roles for myeloid activation^16^, neutrophil activity^17^ and cytotoxic T cell responses^18^ as common features of severe disease. A striking observation in these patients was the complete collapse of germinal centers (GCs) responsible for classical pathways of B cell development in patients that had succumbed to the illness^19^. Nonetheless, patients with severe illness produced high numbers of antibody secreting cells via an alternative extrafollicular (EF) pathway involving the activation of B cell effectors, DN2 B cells, previously described in human autoimmune diseases such as lupus^20,21^. While these responses are highly targeted to the virus, they are also cross reactive against self-antigens resulting in naive B cell-derived, *de novo* autoreactivity^22^. In patients recovering from severe illness with symptoms consistent with PASC, these autoreactive responses were identifiable for months after infection raising the possibility of persistent inflammatory responses continuing to drive symptoms in a subset of patients well into the recovery phase of disease^22^.

Previously, studies of multi-systemic inflammatory syndrome in children (MIS-C) have identified a prolonged systemic inflammatory period following acute infection correlated with innate immune activation, antibody secreting cell expansion, and autoantibody production that is refractory to treatment with immunomodulators^23^. If a subset of patients with ongoing inflammatory activity following COVID-19 recovery similarly exists in adult patients, it follows that they may be uniquely responsive to immunomodulatory therapy. Due to the extraordinarily high disease burden of adult PASC^24^, it is critical to understand if these patient subsets exist within this highly heterogeneous disease and develop tools to accurately identify them.

Here, we show that by making use of refined ML modeling, a subset of PASC patients with ongoing inflammatory processes can be readily identified using serological markers of known biological significance. While more patient follow up is needed, these data represent an important early step in the classification of PASC patients into clinically meaningful subsets.

## Results

### PASC patients display hallmarks of systemic inflammation

To understand the immunologic features underpinning the complex symptomatology associated with PASC, 97 patients were recruited from COVID-19 recovery clinics in Atlanta, GA USA to provide blood samples and deep clinical documentation. Enrollees had a mean age of 50 years (range 21–81), 71 (73%) were female, and the majority were African American (59%) (Table 1). Fifty-seven (59%) had mild acute COVID-19 with the remaining requiring hospitalization. At the time of sampling, patients were a mean of 140 days from COVID-19 onset, with the most common self-reported PASC symptoms included dyspnea (69%), fatigue (64%), and brain fog (47%) (Table 1). Due to inconsistency in formal PASC diagnosis criteria provided by major health organizations for minimum COVID-19 recovery period^13,14^, alongside significant data suggesting that acute-phase disease may predict PASC manifestations^12^, patient samples were collected across a wide range of recovery time points (22-446 DPSO) to understand disease development and potential resolution. Patients who were suspected or diagnosed with rheumatic diseases prior to COVID-19 diagnosis were excluded from the cohort.

**Table 1.** Patient Data Table

| **Characteristics** | **PASC Patients**<br>**(n = 97)** | **Uncomplicated COVID**<br>**recovery (n = 26)** |
| --- | --- | --- |
| **Sex** |  |  |
| Female | 71 (73%) | 10 (50 %) |
| Male | 26 (27%) | 13 (50%) |
| **Age, mean (range)** | 50 (21-81) | 40 (24-70) |
| 20-39 years | 16 (17%) | 10 (67%) |
| 40-59 years | 60 (63%) | 2 (13%) |
| 60-79 years | 18 (19%) | 2 (13%) |
| > 80 years | 1 (1%) | 0 (0%) |
| **Race/Ethnicity** |  |  |
| African American/Black | 57 (59%) | 7 (27%) |
| White | 27 (27%) | 14 (54%) |
| Hispanic | 7 (7%) | 2 (8%) |
| Asian | 2 (2%) | 3 (12%) |
| Unknown | 4 (4%) | 0 (0%) |
| **Acute COVID-19 Severity** |  |  |
| Asymptomatic | 0 | 0 |
| Outpatient | 57 (59%) | 23 (88%) |
| Hospitalized | 37 (39%) | 3 (12%) |
| ICU-admitted | 3 (3%) | 0 |
| **Collection DPSO. Mean (range)** | 140 (22-446) | 110 (18-304) |
| 0-3 months | 39 (41%) | 13 (50%) |
| 3-6 months | 34 (36%) | 8 (31%) |
| 6-12 months | 20 (21%) | 5 (19%) |
| >12 months | 2 (2%)) | 0 (0%) |
| **PASC symptoms (self-reported)** |  |  |
| Dyspnea | 65 (68%) |  |
| Fatigue | 61 (64%) |  |
| Brain Fog | 45 (47%) |  |
| Cough | 31 (33%) |  |
| Headache | 29 (31%) |  |
| Chest Pain | 23 (24%) |  |
| Depression | 20 (21%) |  |
| Myalgias | 19 (20%) |  |
| Weakness | 19 (20%) |  |
| Anxiety | 18 (19%) |  |
| Anosmia/Dysguesia | 15 (16%) |  |
| Arthralgias | 14 (15%) |  |

**Table 2.** Subset Patient Data Table

| **Characteristics** | **<u>inflPASC (n = 49)</u>** | **<u>plaqPASC (n = 48)</u>** |
| --- | --- | --- |
| **Sex** |  |  |
| Female | 34 (70%) | 37 (77 %) |
| Male | 15 (31%) | 11 (23%) |
| **Age, mean (range)** | 53 (32-77) | 48 (24-81) |
| 20-39 years | 7 (14%) | 11 (23%) |
| 40-59 years | 27 (55%) | 32 (67%) |
| 60-79 years | 12 (24%) | 5 (10%) |
| > 80 years | 0 (0%) | 1 (2%) |
| **Race/Ethnicity** |  |  |
| African American/Black | 34 (69%) | 22 (46%) |
| White | 9 (18%) | 19 (40%) |
| Hispanic | 5 (10%) | 2 (4%) |
| Asian | 1 (2%) | 1 (2%) |
| Unknown | 0 (0%) | 4 (8%) |
| **Acute COVID-19 Severity** |  |  |
| Asymptomatic | 0 | 0 |
| Outpatient | 24 (49%) | 33 (69%) |
| Hospitalized | 24 (49%) | 14 (29%) |
| ICU-admitted | 1 (2%) | 1 (2%) |
| **Collection DPSO. Mean (range)** | 123 (24-312) | 156 (22-446) |
| 0-3 months | 24 (49%) | 14 (29%) |
| 3-6 months | 16 (33%) | 19 (40%) |
| 6-12 months | 9 (18%) | 14 (29%) |
| >12 months | 0 (0%) | 1 (2%) |

Due to the critical role that systemic inflammation plays in COVID-19^25^, and early documented associations with PASC^25^, a high-dimensional screen of blood proteomics of almost 3,000 independent targets was performed on patient plasma via the Olink Explore 3072 platform. A matched cohort of 26 donors with uncomplicated recoveries from COVID-19 at similar intervals post symptom onset were included as COVID-recovery (CR) controls (Table 1). Substantial heterogeneity in overall levels of blood markers was observed within the PASC group, with a large fraction of patients showing clear discrimination from the CR cohots based on proteomic signatures, alone (Fig 1a). More than 700 proteins displaying significantly increased abundance in the PASC cohort, with 20 additional proteins significantly decreased in comparison to CR controls (Fig 1b). While elevated protein signatures were diverse in function, many of the most significant hits were inflammatory in nature and have been repeatedly identified as associates of the acute phase of severe COVID-19 including IL-6^26^, IL-8^27^, and NF-kB^28^ (Fig 1b,c).

**Figure 1.**
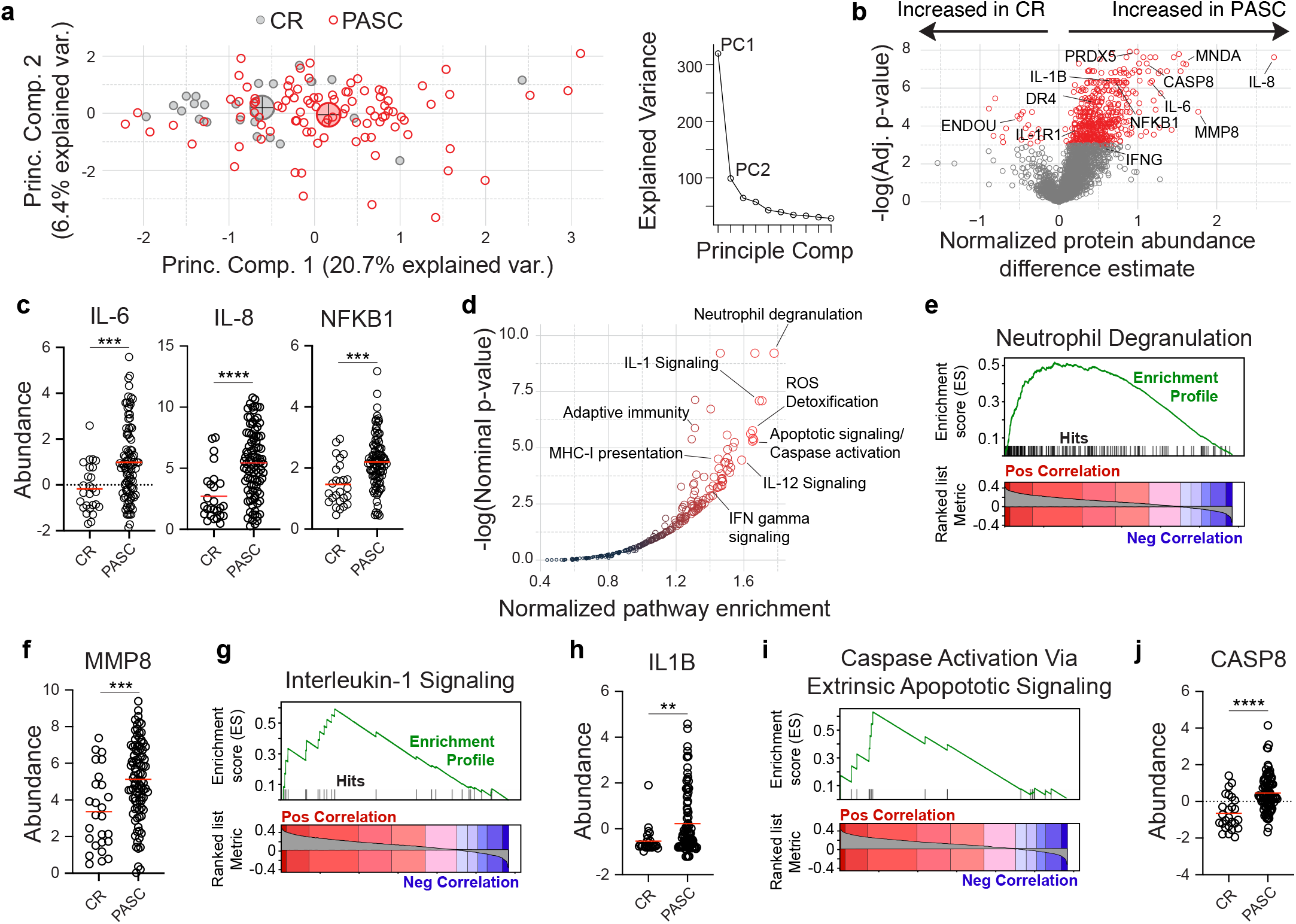
Inflammatory protein signatures in PASC (a-j) Blood plasma from 97 PASC patients and 26 CR controls was assessed for 2925 independent protein features. (a) Left – Principal component analysis of PASC and CR cohorts. Large circles indicate population centroids. Right – Scree plot of the explained sample variance of the first 10 principal components. (b) Feature-wise comparison between PASC and CR cohorts. Proteins of interest are labeled with significant differential abundances (Adj p < 0.001) highlighted in red. (c,f,h,j) Normalized abundance of indicated proteins (d) Reactome pathway analysis of proteins ranked by Spearman’s correlations with PASC diagnosis. Pathways of interest are labeled. (e,g,i) Gene set enrichment analysis of indicated pathways.

To identify broader trends in proteomic alterations within the PASC cohort we identified blocks of related proteins that were enriched in PASC subjects over CR controls (See Methods - Software and analysis). Collectively, an analysis of biological pathway enrichment revealed several interesting biological pathways uniquely associated with the PASC cohort (Fig 1d). Consistent with increased expression of IL-6 and IL-8 (fig 1c) neutrophil degranulation was the most enriched pathway in the set with matrix metalloprotease 8 (MMP8) and myeloid cell nuclear differentiation antigen (MNDA) also highly increased in the PASC cohort (Fig 1b,e,f). While multiple cytokine signaling pathways showed elevation in PASC, the IL-1B pathway was particularly responsive, with elevated levels of both the cytokine itself and primary receptor elevated in the blood (Fig 1b,g,h). The identification of IL-1R1, a transmembrane receptor, within the proteomics screen was reflective of a more generalized and unanticipated ability of this method to identify proteins usually restricted to cellular compartments. Strong increased abundances of markers associated with cell death including caspase 8 and the TNF death receptor, DR4, provide a potential explanation (Fig 1b,i,j), suggesting that increased cellular debris from active cell death may be generally more abundant in these patients.

### ML identifies unanticipated features of PASC

Previous reporting on biological and clinical associations of PASC have yielded mixed results, with some prominent studies finding no clear biologic discrimination between patients with PASC and uncomplicated recovery^29^, while others identify clear distinctions at later time points in disease^30^. To take advantage of the high-dimensional nature of the proteomics dataset, we turned to Random Forests (RF), a class of supervised, non-parametric machine learning (ML) models based on aggregating decision trees. RF models can be trained to take advantage of multiple independent or correlated features to generate probabilistic classifiers for a categorical response variable. (Fig 2a). RF models are particularly well suited for this task, as blocks of co-regulated protein abundances within the dataset suggested that repeated parametric testing underlying feature significance testing may be underpowered (Extended data 1). Further, trained RF models prioritize the ability of a feature to help distinguish between cohorts over measures of statistical deviation, thereby elevating the importance of features that may be less striking when considering only effect size and parametric significance(Fig 1c), but are critical discriminators between cohorts, nonetheless.

**Figure 2.**
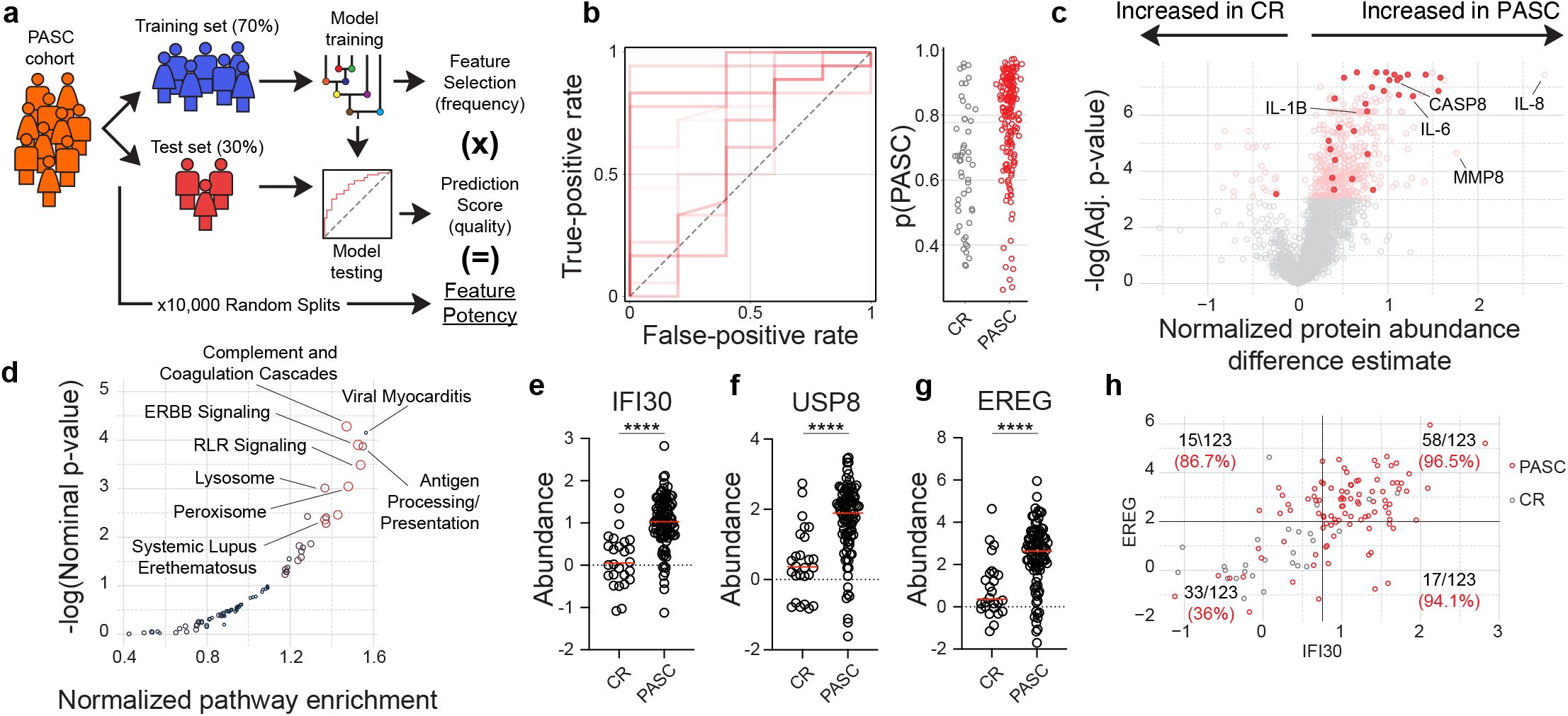
RF modeling identifies PASC features (a-g) Blood plasma from 97 PASC patients and 26 CR controls was assessed for 2925 independent protein features. (a) Cartoon overview of random forest modeling approach and feature potency assessment. (b) Left – Receiver-operating characteristic (ROC) plot displaying 10 models with randomized train/test splits classifying PASC and CR. Right – probabilistic classification plots for individual patients from the test sets derived from the 10 models displayed in the ROC plot (c) Feature-wise comparison between PASC and CR cohorts. Proteins of interest are labeled with the 30 features with highest feature potency highlighted. (d) KEGG pathway analysis of proteins ranked by feature potency in PASC discrimination modeling. (e,f,g) Normalized abundance of indicated proteins. (h) Correlated but distinct information is provided by EREG and IFI30. Fraction of total samples and % purity indicated for each quadrant.

As RF model training is inherently random by design, potentially incorporating sub-optimal features for any individual model, we took a consensus modeling approach whereby 10,000 independent RF models were trained and evaluated on different data splits. This cross validation approach is critical in ensuring that resulting models are not overfit to the dataset and maintain their generalizability to the broader patient population^31^.

Indicating heterogeneity in the PASC patient group, model performance was highly dependent on the patient cohort selected for inclusion within the training set (Fig 2b). To identify individual features associated with strong model performance and PASC generalizability, proteins were individually scored for the frequency of incorporation in a model over 10,000 iterations, the importance of the feature for each model in group discrimination, and the performance of the models it was integrated into (Supplemental table 1). Perhaps unsurprisingly, the most influential features identified in this way were significantly different between the PASC and CR groups, although they were not uniformly the most significant or differentially expressed features in the set (Fig 2c).

Notably, many of the most significantly expressed inflammatory cytokines linked to neutrophil activity, including IL-6 and IL-8, were not identified among the top scoring discriminators of PASC based on blood-based protein profiling (Supplemental table 1). While neutrophil degranulation signatures were highly represented in overall differential expression analyses, an assessment of the biological pathways associated with high feature potency revealed coagulation cascades, endothelial growth factor (EGF) signaling, antiviral sensing, and antigen presentation as effective discriminators of PASC and CR patient groups. In addition, proteins associated with viral myocarditis and SLE were significantly enriched.

These pathways were directly reflected in the most potent individual discriminators of PASC within the feature set. In 10,000 RF models trained on different subsets of subjects, IFI30, an interferon-gamma-induced mediator of peptide processing for MHC loading^32^, was incorporated as a key discriminator of PASC more than 96% of the time and associated with models with high predictive value on held-out data (Fig 2e, Supplemental table 1). USP8, a component of T cell antigen receptor (TCR) signalosome critical for thymocyte development, homeostasis, and proliferation^33^, was incorporated into models with similar frequency, although its selection was less well associated with predictive power (Fig 2f, Supplemental table 1). Perhaps most interestingly, the epidermal growth factor (EGF), epiregulin (EREG), was consistently upregulated in PASC and was selected for incorporation into almost 90% of final predictive models (Fig 2g, Supplemental table 1). EREG has been identified as a critical mediator of IL-6/IL-17-induced upregulation of several EGF members and has been identified as dysregulated across a number of chronic autoimmune conditions^34^. As expected, many of the individual features identified are correlated, but provide unique information for identifying PASC patients (Fig 2h). Overall, application of machine learning to the dataset resulted in the identification of several biological pathways and individual targets with potential value both diagnostically and therapeutically in patients with PASC.

### Broad inflammation defines a subset of PASC

Although RF-based approaches were promising in identifying PASC based on blood proteomics alone, expression of individual markers within the PASC cohort was highly heterogeneous – particularly within protein sets associated with inflammation and neutrophil activity (Fig 1c,f,h). Together with the observation that highly significant markers of inflammation, such as IL-6, were not selected as good discriminators of PASC, this suggested that there may be subsets of the cohort with differential immologic activity signatures. Consistent with this hypothesis, unsupervised clustering of the total recovery cohort into two subsets identified a clear subset of PASC patients clustering together with the CR cohort, while another set segregated almost entirely independently (Fig 3a). Hierarchical clustering of the PASC cohort revealed a clear bifurcation of the overall cohort into two broad subsets (Fig 3b). Assessment of the major markers of inflammation significantly upregulated in PASC such as IL-6, IL-8, and IL-1B all showed significantly increased abundances in one of the two PASC subsets, hereafter referred as the inflammatory PASC (inflPASC) subset (Fig 3c,d). While the other PASC subset -- plasma quiescent PASC (plaqPASC) -- showed elevated levels of many inflammatory cytokines, they often failed to reach significance in reference to the CR cohort (Fig 3d). Indeed, virtually all of the proteins that were significantly increased when comparing CR to PASC are attributable to their differential expression within the inflPASC group (Fig 3e).

**Figure 3.**
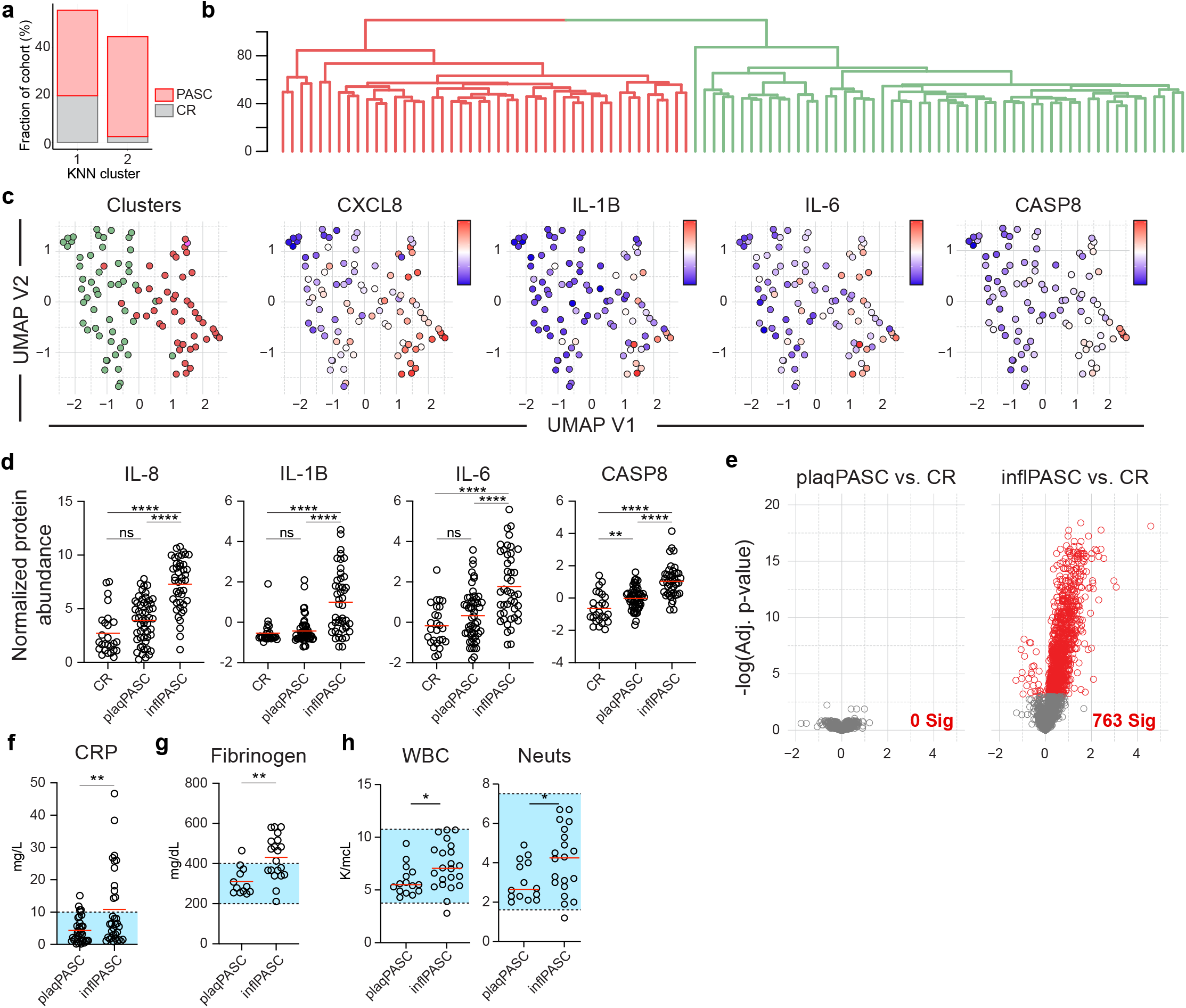
PASC subsets defined by inflammation (a-e) Blood plasma from 97 PASC patients and 26 CR controls was assessed for 2925 independent protein features. (a) Classification of PASC and CR patients following KNN clustering. (b) Hierarchical clustering of PASC patients. Major branches are differentially colored. (c) UMAP analysis of PASC patients displaying cluster classifications from [b], or indicated proteins of interest. (d) Quantification of indicated proteins in CR, plaqPASC, and inflPASC subsets. (e) Feature-wise comparison between CR and either inflPASC or plaqPASC cohorts. (f-h) Clinical blood testing of plaqPASC and inflPASC patient cohorts. Blue boxes indicate normal testing ranges. (f) C-reactive protein concentration in inflPASC and plaqPASC cohorts (g) Fibrinogen in inflPASC and plaqPASC cohorts. (h) White blood cell and neutrophil counts from complete blood counts of inflPASC and plaqPASC patients.

Due to the broad range of time points collected within the recovery cohort, it was possible that the stark separation of PASC patients-based proteomics assessment could simply be attributable to differences in recovery periods between the inflPASC and plaqPASC groups. However, this was not the case. While inflPASC patients trended towards earlier recovery time points, restricting analysis to only those patients with recovery time points exceeding 90 DPSO (thereby exceeding WHO PASC minimum recovery recommendations) continued to show significant differences between plaqPASC and inflPASC cohorts across inflammatory markers (Extended data 2a,b). This, combined with individual observations of inflPASC profiles more than a year post recovery strongly suggest that these responses may be unexpectedly stable in a sizable proportion of patients. In addition to recovery periods, no clear demographic correlate could be identified to singularly explain the identification of the inflPASC disease state. Patient age was slightly higher in the inflPASC group, with the cohort enriched for African American males (Supplementary table 2).

The identification of inflPASC based on clear differences in inflammatory signaling in the blood suggested that this heterogeneity may help explain the mixed results in clinically identifying PASC as a whole through traditional clinical blood testing. This was confirmed as broad markers of inflammation including C-reactive protein (CRP) could be readily identified in the plasma of inflPASC patients versus plaqPASC counterparts (Fig 3f). Clotting factors were also elevated in the inlfPASC group, with significant increases in both fibrinogen (Fig 3g). Further clinical blood testing revealed elevated white blood cell counts in the inflPASC cohort attributable to elevated neutrophil counts corresponding to the neutrophil-focused proteomics signature in these patients (Fig 3h).

### inflPASC patients show active B cell profiles

To understand the nature of the cellular responses underlying the altered humoral targeting in the inflPASC group, targeted flow cytometry was performed on 39 PASC patients (n = 15 inflPASC, 109 DPSO average; n = 23 plaqPASC, 137 DPSO average). In the acute phase of severe COVID-19, extrafollicular B cell responses correlated with the rapid expansion of antibody secreting cells (ASCs)^21^, resulting in both antiviral and anti-self reactivity^22^. Similar B cell profiling of PASC patients revealed a more mixed response. Hierarchical clustering based on B cell subpopulation composition resulted in some aggregation of the inflPASC cohort, however it did not appear dominated by a clear B cell activation pathway; instead reflecting a more general profile of B cell activation in comparison to the plaqPASC group (Fig 4a). This was consistent with elevated profiles of the B cell survival factors BAFF and APRIL in the inflPASC group suggesting ongoing support for B cell activity in the blood (Fig 4b). Of great interest, a positive regulator of activation induced cell death and autoreactive control, Uridylate-Specific Endoribonuclease (EndoU), was decreased in inflPASC patients (Fig 4c).

**Figure 4.**
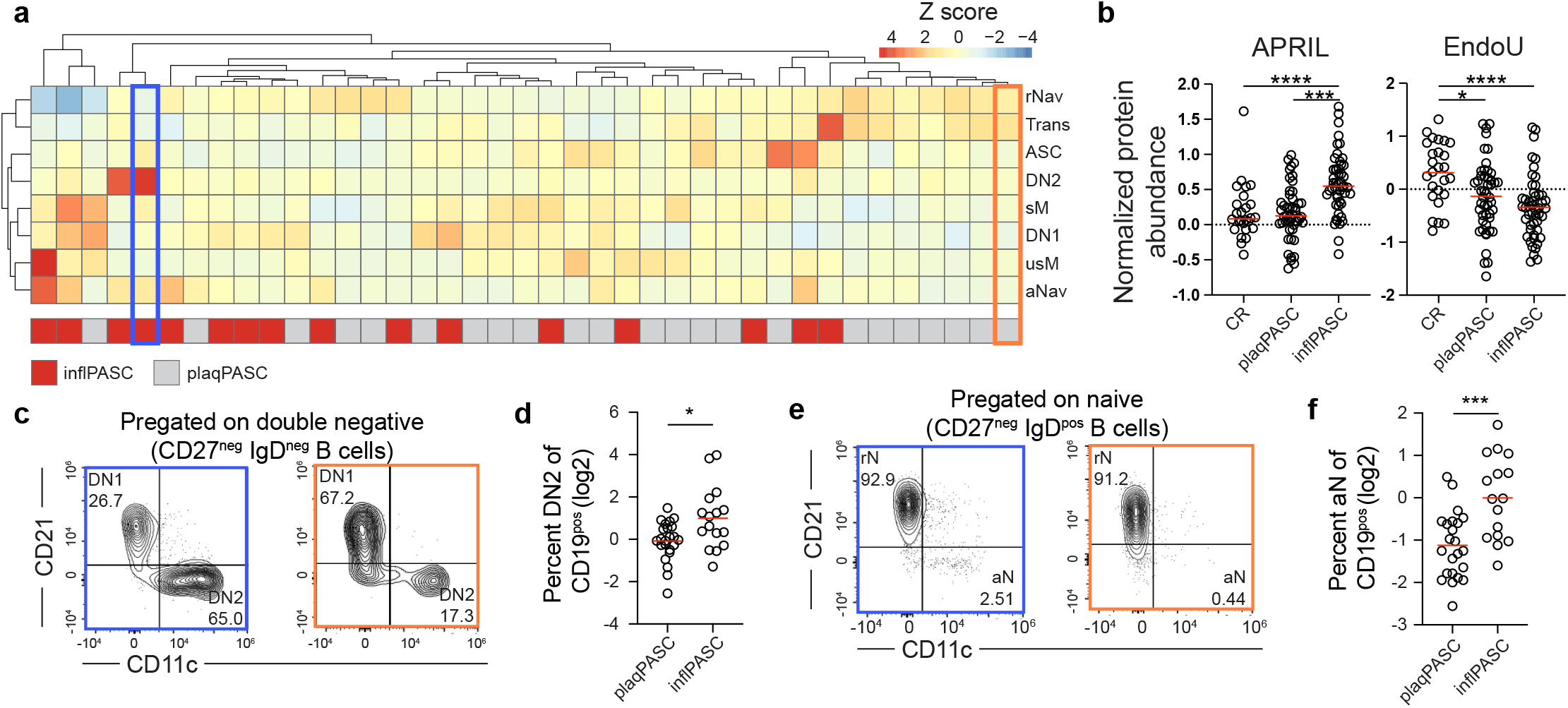
EF B cell activity in inflPASC patients (a,c-g) Flow cytometric analysis B cells from 39 PASC patients. (a) Heat map of B cell population frequency z-scores in inflPASC (red; n = 16) or plaqPASC patients (gray; n = 23). Multivariate clustering of patients by Ward’s method is represented by dendrograms. Colored boxes highlight individual patients for individual flow plot visualizations in [4d] and [4f]. (b) Normalized abundance of indicated proteins (c) Representative plots of DN population composition in inflPASC and plaqPASC patients. (d) DN2 frequency of CD19+ B cells in inflPASC and plaqPASC patients. (e) Representative plots of naive B cell population composition in inflPASC and plaqPASC patients. (f) aN frequency of CD19+ B cells in inflPASC and plaqPASC patients.

While B cell profiling did not reveal the same magnitude of emphasis on the EF B cell pathway as in the acute phase of COVID-19, it was nonetheless enriched within the inflPASC subset. Assessment of the EF effector population, DN2 B cells, showed increased frequencies in the inflPASC group in comparison to the plaqPASC subset (Fig 4c,d). Surprisingly, the presence of activated naive (aN) B cells within the inflPASC cohort was much more pronounced (Fig 4e,f). aN B cells have been previously identified as precursors to the EF response and linked with naive-derived autoreactive antibodies in chronic autoimmune disease^35^. Their presence here, in conjunction with a more muted DN2 effector response, is suggestive of a novel role for these cells beyond the acute phases of an emerging EF-biased response.

### inflPASC patients display altered humoral targeting

In acute COVID-19, high levels of inflammation in critical illness drove higher levels of SARS-CoV-2-targeted antibody responses with significant cross-reactivity against self-antigens^22^. Serological testing of plaqPASC and inflPASC patients identified no clear serological difference between the groups in targeting the SARS-CoV-2 receptor binding domain binding, although IgM and IgA titers were slightly higher in the inflPASC group (Fig 5a). By contrast, non-spike targeting was elevated in inflPASC. In particular, nucleocapsid antibodies were enriched in the inflPASC cohort across all isotypes tested, with significant increases in both IgA and IgG responses (Fig 5b). As anti-nucleocapsid responses are known to diminish significantly over time, it was possible that the differences in targeting were attributable to the established trends in the inflPASC group towards earlierDPSO. However, restricting the analysis to patients collected more than 120 days post-diagnosis and eliminating the early time point bias of the inflPASC group showed similar enrichment of anti-nucleocapsid antibodies, suggesting that these differences in humoral immune targeting are stable over the time periods assessed (Fig 5c).

**Figure 5.**
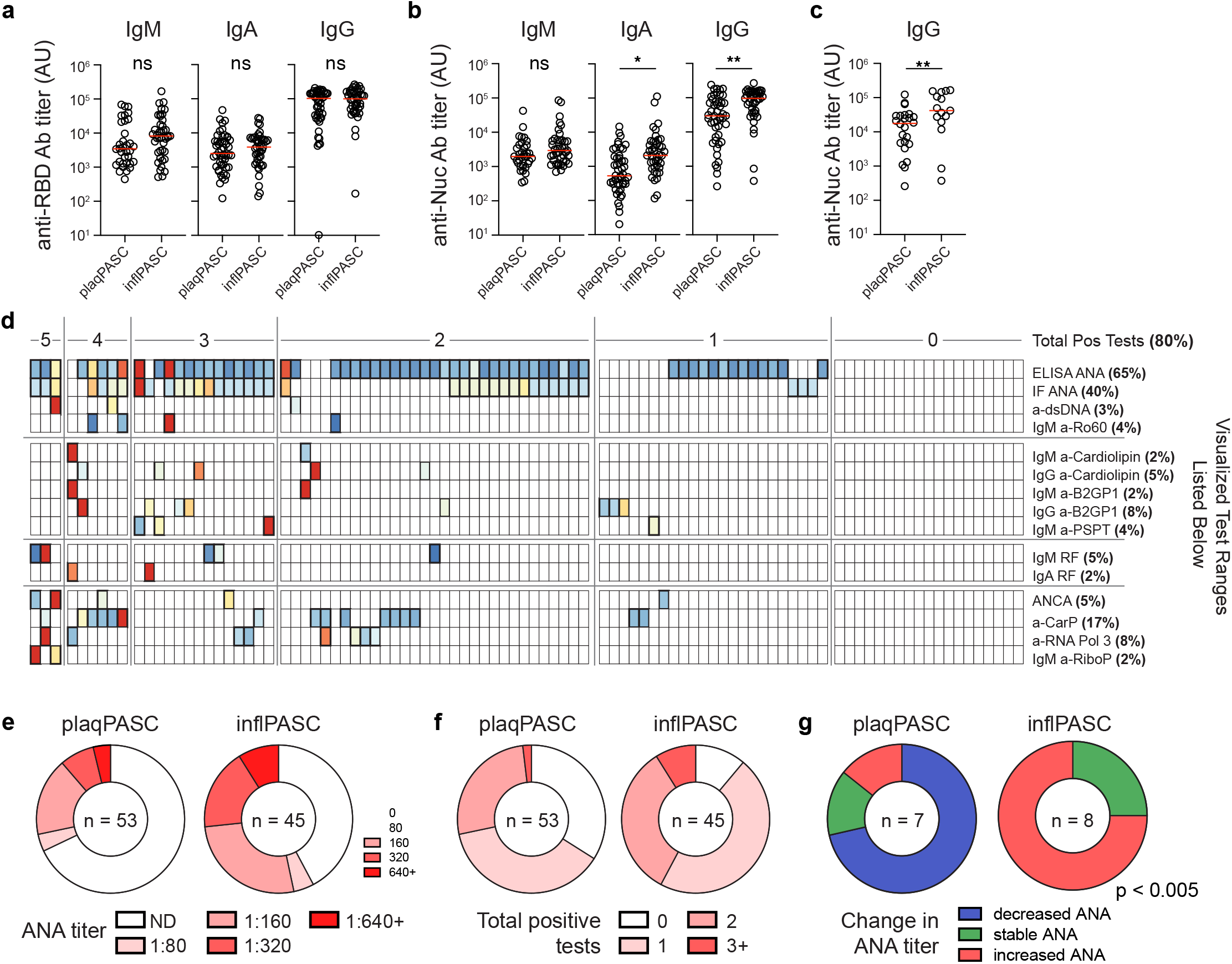
Autoreactive serology in inflPASC (a-c) Plasma from 97 PASC patients was screened for reactivity against SARS-CoV-2. (a) Serological reactivity against the spike receptor binding domain in inflPASC and plaqPASC patients, by isotype. (b) Serological reactivity against nucleocapsid in inflPASC and plaqPASC patients, by isotype. (c) Serological anti-nucleocapsid responses in patients in inflPASC and plaqPASC cohorts more than 120 days DPSO. (d-g) Plasma from 96 PASC patients with mixed symptomology were screened by Exagen clinical laboratory for reactivity against 30 clinically-relevant autoantigens. (a) Heatmap of patient results. Each column represents a single patient grouped by the total number of autoreactive positive tests that the patient displayed. Bolded boxes represent clinical positive tests with the color indicating the magnitude of the test result. Scale for each test is documented below the heatmap. (e) ANA titers in plaqPASC and inflPASC patients. (f) Total positive autoreactive tests in plaqPASC and inflPASC patients. (g) Trend in ANA titers in (n = 7) plaqPASC and (n = 8) inflPASC patients 1 year after initial plasma collection and screening.

Previously, non-spike targeting in multisystem inflammatory syndrome in children (MIS-C) correlated with the production of self-targeted antibodies^23^. This, alongside previous studies in COVID-19 identifying clear autoreactivity profiles in COVID-19 and lingering autoreactivity into the recovery phase suggested that inflPASC patients might also be enriched for autoreactive targeting. To this end, plasma samples were screened against 30 clinically-relevant autoantigens associated with connective tissue disorders. As in acute COVID-19, patients with PASC were enriched for autoreactivity with more than 75% showing reactivity against at least one autoantigen (Fig 5d). Also similar to COVID-19, anti-nuclear antigen (ANA) testing showed broad positivity, although much of the cohort displayed low titers (1:80 - 1:160) of questionable clinical relevance.

However, more than a third of patients displayed autoreactivity against 2 or more autoantigens, with some patients resulting 5 total positive tests (Fig 5d). As in COVID-19, anti-carbamylated protein responses were enriched with 17% of patients testing positive, alongside an unexpected enrichment in RNA polymerase 3 reactivity across the cohort.

While autoreactivity was enriched across the entire PASC cohort, it was further emphasized within the inflPASC subset. As a broad measure of broken tolerance, inflPASC patients displayed both higher incidence (>55%) and higher titers of ANAs. Increased ANA titers were reflective of broader autoreactivity within the group which contained a higher percentage of patients with positive tests to two or more independent self-antigens. Of interest, anti-neutrophil cytoplasmic antibodies (ANCA) were restricted to the inflPASC group (4/44). Further, of the 6 patients resulting positive tests for anti-beta-2 glycoprotein 1 (B2GP1) antibodies, associated with clotting abnormalities in both anti-phospholipid syndrome and COVID-19, 5 segregated into the inflPASC subset.

Critically, a targeted follow up of patients roughly 1 year after initial visit revealed resolving ANA reactivity in plaqPASC patients (5/7) in contrast to the building reactivity in inflPASC patients (6/8) (Fig 5e). Of the eight inflPASC patients with follow up testing, three were initially collected 90+ DPSO and all showed increasing titers demonstrating clear evidence of building autoreactivity beyond the acute phase of COVID-19. Further, one inflPASC patient had developed new reactivity against dsDNA, opening the possibility of antigen walk and chronic autoimmune development.

### Classifying inflPASC through ML

The inflammatory milieu, neutrophilia, discordant self-reactivity and altered B cell responses suggest that the inflPASC cohort may uniquely benefit from immunomodulation in the alleviation of disease burden. To accurately identify this specific patient subset, RF modeling was again implemented, this time classifying inflPASC patients from all other COVID-19 recovery. The resulting predictive modeling was extremely robust – 10,000 models with randomized training/test set splits resulted in a mean ROC AUC of 0.95 (SD +/-0.04) suggesting that, unlike the generalized PASC cohort, inflPASC patients could be efficiently identified irrespective of the patient set selected for model training (Fig 6a). This, combined with the broad set of proteins with increased abundance in inlfPASC patients strongly suggested that restricting our feature set to targets of known immunologic significance might still be effective in parsing the group. To this end, a list of 12 targets was manually curated from the most potent discriminators of inflPASC and used as inputs into a new RF model (Fig 6b). Despite the restricted feature set, use of feature potency scores to guide parameter selection resulted in modeling that continued to be effective in discriminating the inflPASC group with a mean ROC AUC of 0.94 (SD +/-0.05), suggesting that full proteomics screening is not necessary to identify these patients.

**Figure 6.**
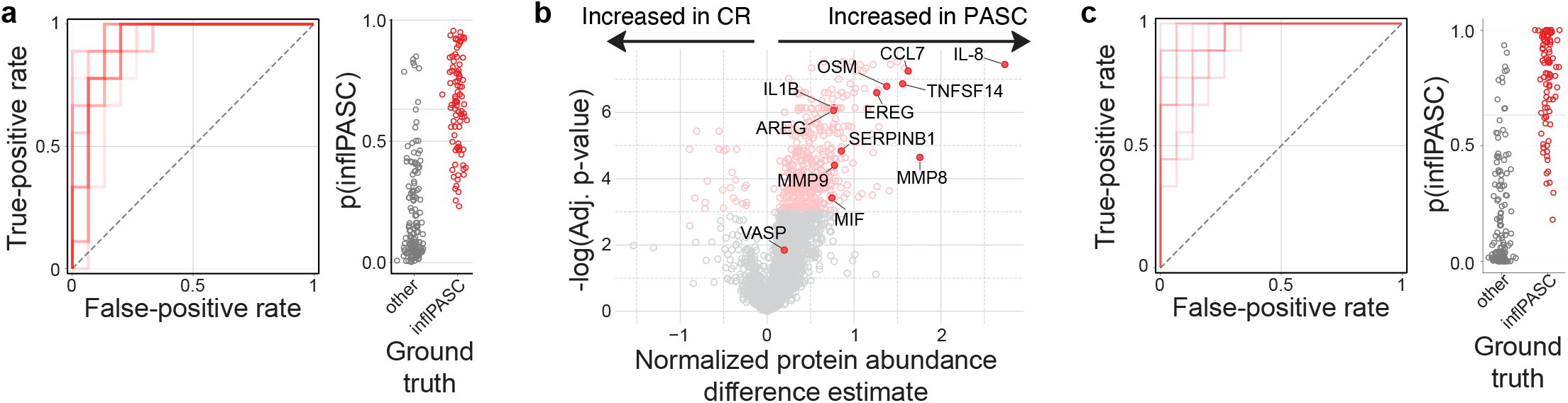
Identifying inflPASC in small feature sets (a) Left – Receiver-operating characteristic (ROC) plot displaying 10 models with randomized train/test splits classifying inflPASC from all other recovery. Right – probability plots for individual ‘test’ patients derived from the 10 models displayed in the ROC plot. (b) Feature-wise comparison between PASC and CR cohorts. 12 manually curated proteins for reduced feature inputs are labeled. (c) Left – Receiver-operating characteristic (ROC) plot displaying 10 models with randomized train/test splits classifying inflPASC from all other recovery using the manually curated list of 12 high-potency proteins identified in [6b]. Right – probability plots for individual ‘test’ patients derived from the 10 models displayed in the ROC plot.

## Discussion

Clinical heterogeneity in patients with PASC has made it challenging to identify clear biological associations with the disease^29^. Here, we suggest that PASC should be subclassified into (at least) two distinct conditions, characterized by the presence (inflPASC) or absence (plaqPASC) of broad inflammatory signatures consistent with high neutrophil activity and ongoing B cell activation. Using high-dimensional proteomics in combination with machine learning-based modeling, we characterize clear signatures of generalized PASC strongly suggestive of dysregulation of discrete biologic processes underlying disease that may be tractable for both diagnostic and therapeutic purposes. While traditional feature-wise testing showed an inflammatory component to PASC as a whole, a finding largely in agreement with emerging literature, pro-inflammatory cytokines such as IL-6, IL-8, and IL-1B were not identified as strong candidates for the discrimination of PASC when modeling the totality of blood protein content. Instead, signatures of complement and clotting cascades, active antigen processing, and EGFR signaling were more consistently associated across the group, with the unanticipated identification of unique targets, such as epiregulin, that may hold important diagnostic value.

Likewise, proteomics-based clustering of PASC patients revealed a clear subset of patients closely associated with inflammatory immune signatures strongly suggestive of neutrophilic activity. Through readily-available clinical testing, these inflPASC patients displayed increased white blood cell counts driven by neutrophil expansion in comparison to plaqPASC counterparts, and showed significant increases in acute-phase reactants such as C-reactive protein, fibrinogen, and D-dimer. Associations with clinically-relevant autoantibodies, particularly those related to autoimmune vascular and clotting disorders, suggest that risk of micro-clotting, now widely reported in patients recovering from COVID-19, may be predictable through accurate identification of PASC subtype. However, these signatures are not readily apparent when taking the PASC cohort as a whole. It is important to note that even while these clinical markers are elevated within the inflPASC group, they do not necessarily reflect ‘abnormal’ test results in all cases. That is, the testing of any marker independently may not, by itself, indicate clear disorder. Instead, the elevation of multiple markers, even when within ‘normal’ ranges, seem to best reflect the broad inflammatory signals identified in the proteomics screen. This finding only emphasizes the need to develop tools capable of providing nuanced assessment across a variety of clinical parameters in patient classification. Similarly, it is important to acknowledge that plaqPASC is defined only as the absence of a clear inflammatory signature in the blood, and not as the absence of disease. As others have now shown^30^, and we show here (Fig 2b,h), biological associations can be readily identified across a wide spectrum of PASC independent of clear inflammatory signaling. It will be critical to understand how all of these signatures predict, and potentially contribute to, long-term patient morbidity.

Cellular immune signatures were correlated with the autoantibody signatures identified in the inflPASC cohort suggesting an increased emphasis of the EF B cell response pathway. Recently, this pathway has been directly implicated in the emergence of autoreactivity in severe COVID-19, with permissive B cell selective pressures leading to the emergence of cross-reactive clones capable of targeting both viral- and self-antigens^22^. However, in contrast with those studies, the prominence of the described effectors of that pathway, DN2 B cells and ASCs, was notably more muted in inflPASC patients. However, the presumed precursor to this pathway, aN B cells, were identified at almost a 2-fold increase over baseline. While more work is needed, these cells consistent with this phenotype have been previously implicated as an antigen-specific memory compartment within the context of yellow fever vaccination^36^. Here, they correlate with persistent reactivity against non-spike SARS-CoV-2 antigens. With previous studies in MIS-C identifying non-structural protein targeting as a feature highly related to autoreactivity, and previous implication of the aN populations in the emergence of de novo autoreactivity in chronic autoimmunity, it will be important to know if these cells may contribute to EF-derived memory responses and/or have a propensity towards self-targeting. Indeed, the identification of increasing autoreactivity in these patients at extended time points after COVID-19 recovery strongly suggests that an autoreactive memory compartment may be identifiable in these patients.

The overwhelming disease burden attributable to PASC worldwide^10^ demands that serious attention must be paid both to its accurate diagnosis as well as potential therapeutic avenues. The identification of a clear subclassification of PASC with a highly inflammatory presentation is an important first step. Based on these data, it is likely that these two PASC subclassifications may respond differently to the immunomodulatory therapies currently being investigated in large-scale clinical trials. Using machine-learning approaches, we have identified critical factors that can be used as positive classifiers of inlfPASC with a high degree of sensitivity and precision. Notably, while initial characterization of this heterogeneity required high-dimensional and unbiased screening, we found that a small subset of features that could be tested at scale, selected through novel assessments of feature potency, was nearly as performant when considered alone. Further, our integration of these data with classical in-clinic blood counts, clotting tests, autoreactive screening, and inflammatory marker assessment suggests that there may be several viable avenues to the positive identification of inflPASC patients without need for highly specialized technology. These assessments could be easily integrated into ongoing clinical trials to understand if therapeutics exert discordant effects on specific patient groups and reduce the potential for false-negative outcomes due to patient heterogeneity.

A critical question building upon these initial findings will be the stability of these inflammatory states over time. A surprising finding from these data, in combination with the published literature^29^, is the difficulty in discriminating disease subtype through symptom presentation alone. While trends do emerge, symptom presentation appears to be a poor discriminator of the inflPASC and plaqPASC groups despite their discordant underlying biology and identification of long-term physiological manifestations. As a result, it will be important to move beyond symptomatic presentation as a method of classifying patients, and understand how the signatures reported here might evolve over the course of disease. To this end, while our cross-sectional approach defines clear lines between plaqPASC and inflPASC, it is not yet clear if these presentations are mutually exclusive. In the case of reservoir-based viral reactivation as a main driver of PASC, as several have argued^37^, it could be that inflPASC manifestations are an observation of an inflammatory phase of cyclic reactivation rather than a discrete patient subtype. However, regardless of the stability of these manifestations, these data, taken together, point to an inflammatory process connected to altered long-term immunologic manifestations that may provide an abundance of therapeutic targets aimed at the alleviation of this debilitating disease.

## Data Availability

Data will be made available, upon request, pending peer-review of the work.

## Methods

### Human subjects and clinical assessment

All research was approved by the Emory University Institutional Review Board (Emory IRB nos. IRB00058507, IRB00057983 and IRB00058271) and was performed in accordance with all relevant guidelines and regulations. Informed consent was obtained from all participants. Donors with uncomplicated COVID-19 recoveries (n = 26) were recruited using promotional materials approved by the Emory University Institutional Review Board.

Patients with PASC (n=97) were referred by primary care providers or by self-referral to Emory University Midtown, Emory University Executive Park, and Grady Memorial Hospital PASC Clinics. Adults aged ≥18 years with documented SARS-CoV-2 antigen or anti-nucleocapsid antibody (64%), or those meeting the CDC COVID-19 clinical case definition who were experiencing new or worsening symptoms and were >14 days from COVID-19 onset (36%) were eligible. Sociodemographic, comorbidity, acute COVID-19, and PASC symptom data were collected by patient report through a review of systems and confirmed through medical record review. Clinical blood testing was performed on a subset of patients through routine care protocols.

Peripheral blood was collected in either heparin sodium tubes (PBMCs) or serum tubes (serum; both BD Diagnostic Systems). Study data were collected and managed using REDCap electronic data capture tools hosted at Emory University.

### Proteomic assessment and analysis

Frozen donor plasma was submitted for analysis using the commercially available Olink Explore 3072 platform. Briefly, individual protein features are targeted by two independent antibodies carrying ssDNA tags. Upon dual-Ab binding, the ssDNA tags hybridize forming a PCR-competent substrate for amplification and sequencing.

Protein abundances are normalized against in-plate and global controls, and reported alongside sensitivity thresholds and quality control metrics. Resulting data was further assessed for quality with 1 PASC patient removed due to generalized protein abundances well outside of normal assay ranges. All samples were generally assessed for normal protein expression distributions, and analyzed either through assessment tools provided by Olink in their custom ‘R’ package, or through customized analysis pipelines developed in-house.

### COVID-19 Multiplex Immunoassay

SARS-CoV-2 antigens were coupled to MagPlex Microspheres of spectrally distinct regions via carbodiimide coupling and tested against patient samples as previously described (2). Results were analyzed on a Luminex FLEXMAP 3D instrument running xPonent 4.3 software. Median fluorescent intensity (MFI) using combined or individual PE-conjugated detection antibodies (anti-IgA/anti-IgG/anti-IgM) was measured using the Luminex xPONENT software on Enhanced PMT setting. The background value of the assay buffer or R10 media was subtracted from the serum/plasma or MENSA results, respectively, to obtain MFI minus background (net MFI). Serum and plasma samples were tested at 1:500 dilution and MENSA was tested undiluted.

### Flow cytometry

Isolated PBMCs (10 × 10^6^) were centrifuged and resuspended in 75 μl FACS buffer (PBS + 2% FBS) and 5 μl Fc receptor block (BioLegend, no. 422302) for 5 min at room temperature. For samples stained with anti-IgG, it was observed that Fc block inappropriately interfered with staining, so a preincubation step of the anti-IgG alone for 5 min at 22 °C was added before the addition of the block. Next, 25 μl of antibody cocktail (Supplementary Table 3) was added (100 μl staining reaction), and samples were incubated for 20 min at 4 °C. Cells were washed in PBS, and resuspended in a PBS dilution of Zombie NIR fixable viability dye (BioLegend, no. 423106). Cells were washed and fixed at 0.8% paraformaldehyde (PFA) for 10 min at 22 °C in the dark before a final wash and resuspension for analysis.

Cells were analyzed on a Cytek Aurora flow cytometer using Cytek SpectroFlo software. Up to 3 × 10^6^ cells were analyzed using FlowJo v10 (Treestar).

### Autoreactivity screening

For autoimmune biomarker analysis, frozen plasma was shipped on dry ice to Exagen, Inc. (Vista, California, USA) which has a clinical laboratory accredited by the College of American Pathologists (CAP) and certified under the Clinical Laboratory Improvement Amendments (CLIA). Thawed plasma was aliquoted and distributed for the following tests: anti-nuclear antibodies (ANA) were measured using enzyme-linked immunosorbent assays (ELISA) (QUANTA Lite; Inova Diagnostics) and indirect immunofluorescence (IFA) (NOVA Lite; Inova Diagnostics); anti-double-stranded DNA (dsDNA) antibodies were also measured by ELISA and were confirmed by IFA with Crithidia luciliae; extractable nuclear antigen autoantibodies (anti-Sm, anti-SS-B/La IgG, anti-Scl-70 IgG, anti-U1RNP IgG, anti-RNP70 IgG, anti-CENP IgG, anti-Jo-1 IgG, and anti-CCP IgG) as well as Rheumatoid Factor (RF) IgA and IgM were measured using the EliA test on the Phadia 250 platform (ThermoFisher Scientific); IgG, IgM, and IgA isotypes of anti-cardiolipin and anti-β2-glycoprotein, as well as anti-Ro52, anti-Ro60, anti-GBM, anti-PR3, and anti-MPO were measured using a chemiluminescence immunoassay (BIO-FLASH; Inova Diagnostics); anti-CarP, anti-RNA-pol-III, and the IgG and IgM isotypes of anti-PS/PT were measured by ELISA (QUANTA Lite; Inova Diagnostics), while C- and P-ANCA were measured by IFA (NOVA Lite; Inova Diagnostics). All assays were performed following the manufacturer’s instructions.

### Software and analysis

Computational analysis was carried out in R (v3.6.2; release 12 Dec 2019). Heat maps were generated using the ‘pheatmap’ library (v1.0.12), with data pre normalized (log-transformed z-scores calculated per feature) before plotting. Clustering was carried out using Ward’s method. Custom plotting, such as biological pathway analysis, was performed using the ‘ggplot2’ library for base analysis, and then post-processed in Adobe Illustrator. UMAP coordinates were generated using the ‘UMAP’ library, and then visualized through the ‘ggplot2’ library package. GSEA analyses were performed using the GSEA desktop application using Reactome or KEGG gene sets. Statistical analyses were performed directly in R, or in GraphPad Prism (v8.2.1).

### Patient classification through machine learning

Random forest models were trained using ‘MLJ.jl’ and ‘DecisionTrees.jl’. Hyperparameter tuning (maximum splits, minimum number of samples to allow split, minimum number of samples per leaf) for each class of models (CR vs PASC, inflPASC vs Other) was performed independently using a subset of 80% of samples. Iterative training was performed as follows:

1. A stable random number generator seed was selected
2. Samples were randomly assigned to training (80%) and test (20%) sets
3. The model was trained on the training set using 1000 trees, and hyperparameters identified from tuning step
4. Gini (impurity) feature importance was calculated from training data
5. AUC for the model was calculated based on classifications of the test set.
6. Importance scoring for feature $f$ and model $M$ was calculated as $Score(f|M) = Gini(f) * AUC(M)$

## Figure Legends

**Extended data 1.**
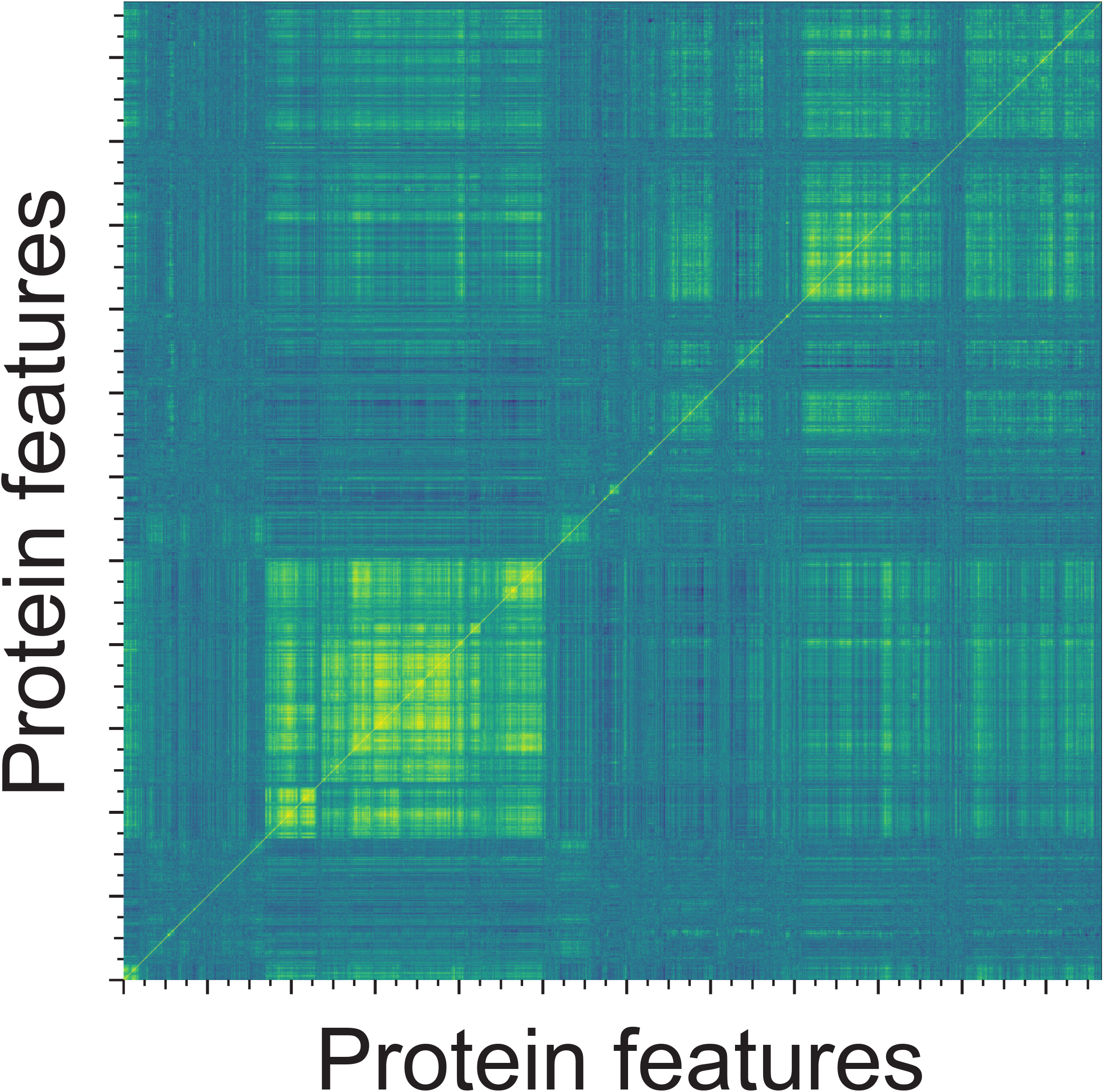
Correlation matrix NxN matrix of total protein features representing large blocks of correlation structure in the protein feature dataset

**Extended data 2.**
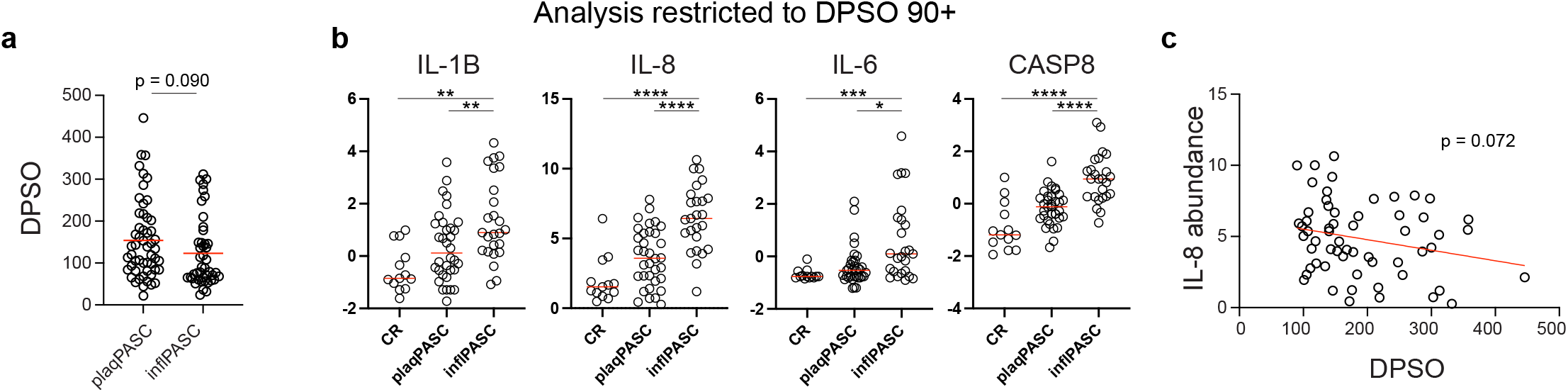
inflPASC designations are not DPSO dependent (a) DPSO of plaqPASC and inflPASC patients. (b) Normalized abundance of indicated proteins of plaqPASC and inflPASC patients with collected at or exceeding 90 DPSO. (c) IL-8 abundance as a function of DSPO in PASC patients.

## Author Contributions

MCW, KSB, SAJ, FEL, and IS conceived of and directed this study. FAM and NSH performed serological evaluation of patient plasma against viral antigens. TAW, MCR, ADT, and AND provided critical patient samples for the study. TAW, CYK, MCR, RPR, and AK conducted chart review and identified samples for study inclusion. YI performed flow cytometric assessment of patient samples. MER oversaw collaborations in autoreactivity testing. MCW and KSB analyzed and compiled all data related to the manuscript. MCW, KSB, and IS wrote the manuscript with all authors providing editorial support.

---

**Supplemental table 1.** PASC vs uncomplicated recovery feature importance list

| gene | sum | count | score |
| --- | --- | --- | --- |
| IFI30 | 87.003465 | 9595 | 69.6045633 |
| USP8 | 52.2644861 | 9635 | 41.6607934 |
| EREG | 50.1070824 | 9056 | 40.0449466 |
| PRDX5 | 48.4315264 | 8932 | 38.5306567 |
| MNDA | 41.5225941 | 8627 | 33.1614331 |
| LMNB1 | 38.2087731 | 8519 | 30.3309411 |
| NPM1 | 37.8243787 | 8594 | 29.8931511 |
| MASP1 | 36.8061305 | 8632 | 29.6699949 |
| CASP8 | 36.9543816 | 8469 | 29.5028407 |
| TRIM21 | 34.8661691 | 8325 | 28.1910787 |
| STX8 | 31.9619348 | 8147 | 25.7162969 |
| APEX1 | 30.8385723 | 7974 | 24.4037579 |
| TOP2B | 29.45735 | 7889 | 23.3982176 |
| KEL | 29.1982307 | 8097 | 23.3026199 |
| SMNDC1 | 29.133717 | 7944 | 23.0904371 |
| RAB44 | 29.0772852 | 7862 | 23.0623292 |
| CC2D1A | 29.2514804 | 7842 | 22.9900151 |
| ELOA | 29.0033986 | 7925 | 22.8746791 |
| CFD | 27.4236633 | 7807 | 22.8149067 |
| PRDX1 | 28.2506547 | 7762 | 22.5065932 |
| GPD1 | 27.8536964 | 7893 | 22.4727361 |
| SNRPB2 | 28.2689169 | 7905 | 22.2635069 |
| CD70 | 25.9360989 | 7744 | 20.9413431 |
| LGALS3 | 25.0963415 | 7629 | 20.49312 |
| CTSS | 25.6233675 | 7676 | 20.1901991 |
| CD300E | 24.5933341 | 7649 | 19.6629619 |
| NUDT16 | 24.7619263 | 7446 | 19.6579643 |
| VTA1 | 24.3946659 | 7500 | 19.5081987 |
| LACTB2 | 24.0078133 | 7432 | 19.0312558 |
| MAD1L1 | 24.0279141 | 7682 | 18.9793636 |
| HNRNPUL1 | 23.8728643 | 7454 | 18.8452374 |
| VSIR | 23.5219994 | 7583 | 18.4109338 |
| CLC | 22.9341129 | 7370 | 18.3962461 |
| RALB | 23.0122683 | 7361 | 18.3161435 |
| C2CD2L | 22.8474453 | 7405 | 18.0861594 |
| NHLRC3 | 22.553694 | 7467 | 17.9546594 |
| LGALS8 | 22.6152379 | 7437 | 17.8509123 |
| F10 | 21.8864242 | 7626 | 17.804826 |
| ABHD14B | 20.9035525 | 7145 | 16.6012046 |
| BACH1 | 20.5193001 | 7319 | 16.5800053 |
| HAVCR2 | 20.8012212 | 7194 | 16.5785581 |
| CXCL8 | 20.6639974 | 7144 | 16.4654265 |
| PGD | 20.6907049 | 7034 | 16.3882535 |
| SCT | 19.2482083 | 7162 | 15.7742875 |
| BRD3 | 19.4607662 | 6988 | 15.7509409 |
| IQGAP2 | 19.6026947 | 7112 | 15.6225802 |
| SAMD9L | 19.7786606 | 6779 | 15.597663 |
| PLAUR | 19.0270611 | 6924 | 15.1109727 |
| RWDD1 | 19.1625383 | 6901 | 15.0943673 |
| LGALS9 | 18.7255874 | 6799 | 14.9358407 |
| SORT1 | 18.3497017 | 7010 | 14.9311718 |
| OPLAH | 17.5452472 | 6820 | 14.3200712 |
| NAPRT | 17.3902881 | 6792 | 14.1045724 |
| ENO1 | 17.4996061 | 6591 | 13.8684762 |
| SERPINB1 | 17.5479911 | 6587 | 13.8239932 |
| NADK | 17.4738172 | 6552 | 13.8153307 |
| PAG1 | 17.5674339 | 7048 | 13.7770855 |
| DFFA | 17.1609117 | 6688 | 13.7675631 |
| PAMR1 | 17.0072864 | 7012 | 13.7572897 |
| MPIG6B | 17.4271994 | 6891 | 13.7480758 |
| FES | 16.5473872 | 6900 | 13.7411702 |
| RAB6A | 16.9345058 | 6694 | 13.5678253 |
| ALDH2 | 16.2818043 | 6729 | 13.47217 |
| ACP6 | 16.3162401 | 6765 | 13.387432 |
| CELA2A | 16.1333799 | 6962 | 13.3825035 |
| GPI | 16.9855032 | 6489 | 13.3453189 |
| TBC1D5 | 16.4788865 | 6699 | 13.2219873 |
| ESM1 | 16.2271801 | 6662 | 13.1260484 |
| SEPTIN8 | 16.3817523 | 6415 | 13.0781986 |
| FIS1 | 16.2261925 | 6581 | 12.8721262 |
| HEPH | 15.8617956 | 6668 | 12.7537828 |
| ENDOU | 15.4213448 | 6695 | 12.730853 |
| CIAPIN1 | 16.2584095 | 6329 | 12.6930769 |
| FAS | 15.5657336 | 6448 | 12.6449128 |
| TSHB | 15.0820913 | 6703 | 12.3917441 |
| METAP2 | 15.7613485 | 6319 | 12.3792753 |
| ASAH1 | 15.5849305 | 6415 | 12.3537836 |
| F2R | 15.3700598 | 6357 | 12.3147351 |
| IST1 | 15.4209877 | 6253 | 12.205614 |
| MYCBP2 | 15.2350225 | 6480 | 12.1745206 |
| MEPE | 15.0450935 | 6496 | 12.1236793 |
| RNF41 | 15.335902 | 6529 | 12.1090473 |
| SCARF1 | 15.0240452 | 6652 | 12.102648 |
| DAG1 | 15.3632509 | 6132 | 12.0441254 |
| GCHFR | 14.8439276 | 6343 | 11.9020907 |
| PIGR | 14.4724914 | 6532 | 11.821503 |
| DGKA | 14.6066732 | 6373 | 11.6657391 |
| GP5 | 14.2914468 | 6685 | 11.6492153 |
| DUSP13 | 14.1986208 | 6416 | 11.5674066 |
| TXLNA | 14.6022945 | 6114 | 11.4858148 |
| HCLS1 | 14.482421 | 6097 | 11.3704339 |
| MKI67 | 14.0938757 | 6584 | 11.3306265 |
| POMC | 13.7132598 | 6388 | 11.0883047 |
| VTI1A | 13.9564038 | 5956 | 11.0053829 |
| AREG | 13.5118595 | 6273 | 10.9364075 |
| CSF1 | 13.4203334 | 6076 | 10.7365721 |
| FEN1 | 13.0835092 | 6160 | 10.6446157 |
| INPP5D | 13.4167507 | 6081 | 10.6435787 |
| APBB1IP | 13.3921463 | 5887 | 10.5384223 |
| SIRPB1 | 12.9653842 | 6402 | 10.4122611 |
| GIGYF2 | 12.8600212 | 5938 | 10.3122015 |
| LPCAT2 | 12.9540109 | 5920 | 10.2264327 |
| GNAS | 12.3713202 | 6452 | 10.1527215 |
| MTPN | 12.3290966 | 7346 | 10.1194995 |
| CEACAM3 | 12.1981619 | 6092 | 9.99937527 |
| PPP1R12A | 12.3521611 | 5722 | 9.95281659 |
| C3 | 12.3776256 | 6091 | 9.90905439 |
| RIDA | 12.101038 | 5956 | 9.82638102 |
| DNAJA2 | 12.5443276 | 5694 | 9.81698236 |
| RABEP1 | 12.433341 | 5700 | 9.79438604 |
| HIP1R | 12.3558728 | 5742 | 9.75204866 |
| F11 | 11.8003271 | 5960 | 9.66930288 |
| CST6 | 11.8101969 | 5955 | 9.66798781 |
| PSPN | 11.5818375 | 6181 | 9.58969631 |
| GP6 | 12.2047854 | 5856 | 9.58385493 |
| ALCAM | 11.7221411 | 6021 | 9.48895833 |
| PSIP1 | 11.7792125 | 5770 | 9.4738083 |
| HSPA1A | 11.9902845 | 5703 | 9.44714574 |
| STX7 | 11.8433787 | 5896 | 9.42958121 |
| TNFRSF1B | 11.6761099 | 5910 | 9.34321221 |
| CRADD | 11.8822958 | 5536 | 9.33999578 |
| MZB1 | 11.6194337 | 5711 | 9.29062169 |
| SELP | 11.5182563 | 5739 | 9.23810354 |
| BID | 11.4133509 | 5444 | 9.09882749 |
| TNFSF14 | 11.3997095 | 5465 | 9.08939936 |
| CA13 | 11.4857975 | 5654 | 9.07544834 |
| PCBD1 | 11.1820159 | 5832 | 9.06776213 |
| COL9A1 | 11.0875779 | 6211 | 9.05136801 |
| TGFA | 11.3468636 | 5550 | 9.03954839 |
| UBE2L6 | 11.3152031 | 5609 | 8.91799911 |
| CEP20 | 11.1862512 | 5773 | 8.89829857 |
| CD46 | 11.3162915 | 5660 | 8.88965213 |
| SERPINB6 | 11.2703913 | 5414 | 8.85370744 |
| TBCB | 11.1744501 | 5397 | 8.8477233 |
| SERPINB9 | 11.1123783 | 5737 | 8.83319676 |
| SLAMF8 | 10.808068 | 5673 | 8.77678491 |
| CRISP2 | 10.5243525 | 5839 | 8.76855614 |
| SCARF2 | 10.7444272 | 5912 | 8.7474878 |
| SELE | 10.7077843 | 5784 | 8.71123195 |
| OSM | 10.8582908 | 5609 | 8.68014757 |
| AHCY | 10.8837037 | 5378 | 8.67134974 |
| GGA1 | 10.5101056 | 6088 | 8.66918301 |
| VIM | 10.8047 | 6294 | 8.66263062 |
| TLR3 | 10.5691445 | 5936 | 8.52003234 |
| S100A11 | 10.8158668 | 5553 | 8.4938598 |
| CSTB | 10.654217 | 5550 | 8.48309145 |
| DCTN2 | 10.7173582 | 5406 | 8.47779015 |
| DMP1 | 10.4993194 | 5704 | 8.46309132 |
| UFD1 | 10.4395178 | 5762 | 8.39926226 |
| B3GNT7 | 10.4355251 | 5723 | 8.38462736 |
| HGF | 10.5150412 | 5339 | 8.38403413 |
| LSP1 | 10.628788 | 5501 | 8.3395892 |
| FYB1 | 10.4413796 | 5318 | 8.33624517 |
| A1BG | 10.2814179 | 5406 | 8.27453606 |
| BRAP | 10.3637938 | 5084 | 8.18066526 |
| DNMBP | 10.3379448 | 5334 | 8.14652068 |
| INSL3 | 9.91769319 | 5811 | 8.1242655 |
| BTN1A1 | 9.85324172 | 5886 | 8.11504713 |
| UGDH | 9.67294622 | 5768 | 8.06819316 |
| SCAMP3 | 10.1093773 | 5307 | 8.02126177 |
| SSC4D | 9.59846371 | 5678 | 8.00785871 |
| SORD | 9.95734322 | 5454 | 7.99013705 |
| SAFB2 | 9.77726857 | 5409 | 7.9444705 |
| GTF2IRD1 | 9.54882524 | 5852 | 7.85279713 |
| LBR | 10.0627606 | 4947 | 7.83269295 |
| NSFL1C | 9.95780713 | 5066 | 7.8258736 |
| CELSR2 | 9.50512397 | 5618 | 7.75370056 |
| PDLIM7 | 9.88691821 | 5053 | 7.73545575 |
| MED21 | 9.38221999 | 5722 | 7.71789418 |
| GOPC | 9.42487145 | 5146 | 7.63358703 |
| SH3BP1 | 9.54808412 | 5351 | 7.63046553 |
| IL1RN | 9.58793885 | 5300 | 7.62554591 |
| MYOC | 9.28064719 | 5748 | 7.62057044 |
| EVI5 | 9.52224164 | 5146 | 7.60741978 |
| ENO2 | 9.60352398 | 5107 | 7.60133757 |
| BIN2 | 9.59939334 | 5134 | 7.58950123 |
| MAP2K1 | 9.62234708 | 4862 | 7.58261399 |
| SCARB2 | 9.48831681 | 5134 | 7.58247072 |
| AP3B1 | 9.48912808 | 5064 | 7.49904605 |
| TYMP | 9.52556685 | 5252 | 7.48427698 |
| LEFTY2 | 9.34521978 | 5873 | 7.47579283 |
| TSNAX | 9.02839503 | 5695 | 7.46186358 |
| SIGLEC1 | 9.44102176 | 5345 | 7.44165706 |
| DSG3 | 8.87100451 | 5510 | 7.4064231 |
| CASP2 | 9.33754544 | 5013 | 7.38852023 |
| EIF4G1 | 9.28681846 | 4997 | 7.32755682 |
| TCN1 | 9.20621226 | 5053 | 7.32680357 |
| TNFRSF21 | 8.97483328 | 5612 | 7.29762856 |
| LRG1 | 8.84221541 | 5522 | 7.25692432 |
| GASK1A | 8.93387528 | 5336 | 7.22810116 |
| STX4 | 9.10266564 | 5048 | 7.20820534 |
| SPART | 9.1849463 | 4901 | 7.18488245 |
| HAVCR1 | 8.89888732 | 5223 | 7.15169411 |
| SFRP1 | 8.95785447 | 5168 | 7.14940654 |
| NFKB1 | 8.87600668 | 4966 | 7.08682752 |
| SHBG | 8.57576401 | 5687 | 7.07383505 |
| FCGR2B | 8.60064317 | 5686 | 7.05450414 |
| DDX58 | 8.79940484 | 4850 | 7.02908268 |
| VWC2L | 8.59243352 | 5445 | 7.0250431 |
| S100P | 8.78117364 | 5636 | 7.0195332 |
| EZR | 8.84969523 | 4997 | 6.99968777 |
| QSOX1 | 8.66565031 | 4862 | 6.99580091 |
| KLK4 | 8.44741131 | 5316 | 6.88493199 |
| SPINK5 | 8.27374522 | 5386 | 6.82243661 |
| BCL2L15 | 8.62790308 | 4796 | 6.82241546 |
| AIF1 | 8.45402906 | 4991 | 6.81528463 |
| SEPTIN9 | 8.53768505 | 5027 | 6.80617545 |
| ANXA5 | 8.21345171 | 5279 | 6.78743213 |
| NAGPA | 8.26313301 | 5432 | 6.77825725 |
| CTSL | 8.37909864 | 5028 | 6.75317244 |
| PRL | 8.36383468 | 5393 | 6.73560904 |
| USP28 | 8.53920955 | 4802 | 6.73433901 |
| CD82 | 8.04495416 | 5323 | 6.73209033 |
| LRRFIP1 | 8.61608998 | 4727 | 6.73019101 |
| SLC9A3R1 | 8.15883834 | 5037 | 6.61775715 |
| CWC15 | 8.36117309 | 4731 | 6.61528877 |
| CRACR2A | 8.45926525 | 4913 | 6.61282236 |
| GFRA2 | 8.19118917 | 5201 | 6.59032575 |
| DBI | 8.43308006 | 4524 | 6.58802518 |
| ATRAID | 8.16834313 | 5013 | 6.5792952 |
| ADAM8 | 8.35938231 | 4567 | 6.55885717 |
| CD164 | 8.29914527 | 4860 | 6.53548734 |
| CAMLG | 8.00276809 | 5380 | 6.51608634 |

## References

1 Brodin, P. Immune determinants of COVID-19 disease presentation and severity. Nat Med 27, 28–33, doi:10.1038/s41591-020-01202-8 (2021).

2 Zhang, Q., Bastard, P., Effort, C. H. G., Cobat, A. & Casanova, J. L. Human genetic and immunological determinants of critical COVID-19 pneumonia. Nature 603, 587–598, doi:10.1038/s41586-022-04447-0 (2022).

3 Merad, M., Blish, C. A., Sallusto, F. & Iwasaki, A. The immunology and immunopathology of COVID-19. Science 375, 1122–1127, doi:10.1126/science.abm8108 (2022).

4 Narasimhan, H., Wu, Y., Goplen, N. P. & Sun, J. Immune determinants of chronic sequelae after respiratory viral infection. Sci Immunol 7, eabm7996, doi:10.1126/sciimmunol.abm7996 (2022).

5 Proal, A. D. & VanElzakker, M. B. Long COVID or Post-acute Sequelae of COVID-19 (PASC): An Overview of Biological Factors That May Contribute to Persistent Symptoms. Front Microbiol 12, 698169, doi:10.3389/fmicb.2021.698169 (2021).

6 Munipalli, B., Seim, L., Dawson, N. L., Knight, D. & Dabrh, A. M. A. Post-acute sequelae of COVID-19 (PASC): a meta-narrative review of pathophysiology, prevalence, and management. SN Compr Clin Med 4, 90, doi:10.1007/s42399-022-01167-4 (2022).

7 Gandhi, R. T., Lynch, J. B. & Del Rio, C. Mild or Moderate Covid-19. N Engl J Med 383, 1757–1766, doi:10.1056/NEJMcp2009249 (2020).

8 Siordia, J. A., Jr. Epidemiology and clinical features of COVID-19: A review of current literature. J Clin Virol 127, 104357, doi:10.1016/j.jcv.2020.104357 (2020).

9 Lopez-Leon, S. et al. More than 50 long-term effects of COVID-19: a systematic review and meta-analysis. Sci Rep 11, 16144, doi:10.1038/s41598-021-95565-8 (2021).

10 Poudel, A. N. et al. Impact of Covid-19 on health-related quality of life of patients: A structured review. PLoS One 16, e0259164, doi:10.1371/journal.pone.0259164 (2021).

11 Helmsdal, G. et al. Long COVID in the Long Run-23-Month Follow-up Study of Persistent Symptoms. Open Forum Infect Dis 9, ofac270, doi:10.1093/ofid/ofac270 (2022).

12 Su, Y. et al. Multiple early factors anticipate post-acute COVID-19 sequelae. Cell 185, 881–895 e820, doi:10.1016/j.cell.2022.01.014 (2022).

13 Control, C. f. D. Post-COVID Conditions: Information for Healthcare Providers. (2022).

14 Organization, W. H. Coronavirus disease (COVID-19): Post COVID-19 condition. (2021).

15 Mathew, D. et al. Deep immune profiling of COVID-19 patients reveals distinct immunotypes with therapeutic implications. Science 369, doi:10.1126/science.abc8511 (2020).

16 Arunachalam, P. S. et al. Systems biological assessment of immunity to mild versus severe COVID-19 infection in humans. Science 369, 1210–1220, doi:10.1126/science.abc6261 (2020).

17 Ackermann, M. et al. Patients with COVID-19: in the dark-NETs of neutrophils. Cell Death Differ 28, 3125–3139, doi:10.1038/s41418-021-00805-z (2021).

18 Bange, E. M. et al. CD8(+) T cells contribute to survival in patients with COVID-19 and hematologic cancer. Nat Med 27, 1280–1289, doi:10.1038/s41591-021-01386-7 (2021).

19 Kaneko, N. et al. The Loss of Bcl-6 Expressing T Follicular Helper Cells and the Absence of Germinal Centers in COVID-19. SSRN, 3652322, doi:10.2139/ssrn.3652322 (2020).

20 Jenks, S. A. et al. Distinct Effector B Cells Induced by Unregulated Toll-like Receptor 7 Contribute to Pathogenic Responses in Systemic Lupus Erythematosus. Immunity 49, 725–739 e726, doi:10.1016/j.immuni.2018.08.015 (2018).

21 Woodruff, M. C. et al. Extrafollicular B cell responses correlate with neutralizing antibodies and morbidity in COVID-19. Nat Immunol 21, 1506–1516, doi:10.1038/s41590-020-00814-z (2020).

22 Woodruff, M. C. et al. Dysregulated naive B cells and de novo autoreactivity in severe COVID-19. Nature, doi:10.1038/s41586-022-05273-0 (2022).

23 Ramaswamy, A. et al. Immune dysregulation and autoreactivity correlate with disease severity in SARS-CoV-2-associated multisystem inflammatory syndrome in children. Immunity 54, 1083–1095 e1087, doi:10.1016/j.immuni.2021.04.003 (2021).

24 Xie, Y., Bowe, B. & Al-Aly, Z. Burdens of post-acute sequelae of COVID-19 by severity of acute infection, demographics and health status. Nat Commun 12, 6571, doi:10.1038/s41467-021-26513-3 (2021).

25 Merad, M. & Martin, J. C. Pathological inflammation in patients with COVID-19: a key role for monocytes and macrophages. Nat Rev Immunol 20, 355–362, doi:10.1038/s41577-020-0331-4 (2020).

26 Chen, L. Y. C., Hoiland, R. L., Stukas, S., Wellington, C. L. & Sekhon, M. S. Confronting the controversy: interleukin-6 and the COVID-19 cytokine storm syndrome. Eur Respir J 56, doi:10.1183/13993003.03006-2020 (2020).

27 Li, L. et al. Interleukin-8 as a Biomarker for Disease Prognosis of Coronavirus Disease-2019 Patients. Front Immunol 11, 602395, doi:10.3389/fimmu.2020.602395 (2020).

28 Kircheis, R. et al. NF-kappaB Pathway as a Potential Target for Treatment of Critical Stage COVID-19 Patients. Front Immunol 11, 598444, doi:10.3389/fimmu.2020.598444 (2020).

29 Sneller, M. C. et al. A Longitudinal Study of COVID-19 Sequelae and Immunity: Baseline Findings. Ann Intern Med 175, 969–979, doi:10.7326/M21-4905 (2022).

30 Klein, J. et al. Distinguishing features of Long COVID identified through immune profiling. medRxiv, doi:10.1101/2022.08.09.22278592 (2022).

31 Volovici, V., Syn, N. L., Ercole, A., Zhao, J. J. & Liu, N. Steps to avoid overuse and misuse of machine learning in clinical research. Nat Med, doi:10.1038/s41591-022-01961-6 (2022).

32 Dudziak, D. et al. Differential antigen processing by dendritic cell subsets in vivo. Science 315, 107–111, doi:10.1126/science.1136080 (2007).

33 Dufner, A. et al. The ubiquitin-specific protease USP8 is critical for the development and homeostasis of T cells. Nat Immunol 16, 950–960, doi:10.1038/ni.3230 (2015).

34 Harada, M. et al. Temporal expression of growth factors triggered by epiregulin regulates inflammation development. J Immunol 194, 1039–1046, doi:10.4049/jimmunol.1400562 (2015).

35 Tipton, C. M. et al. Diversity, cellular origin and autoreactivity of antibody-secreting cell population expansions in acute systemic lupus erythematosus. Nat Immunol 16, 755–765, doi:10.1038/ni.3175 (2015).

36 Wec, A. Z. et al. Longitudinal dynamics of the human B cell response to the yellow fever 17D vaccine. Proc Natl Acad Sci U S A 117, 6675–6685, doi:10.1073/pnas.1921388117 (2020).

37 Swank, Z. et al. Persistent circulating SARS-CoV-2 spike is associated with post-acute COVID-19 sequelae. Clin Infect Dis, doi:10.1093/cid/ciac722 (2022).

